# Characterizing Autism Spectrum Disorder in the All of Us Research Program

**DOI:** 10.1101/2025.09.10.25335344

**Authors:** Danni Liu, Yan Li, Zhongli Jiang, Peter J. Chung, Jean-G. Gehricke, Dabao Zhang, Min Zhang

## Abstract

Many autism spectrum disorder (ASD) studies have been established for early diagnosis in youth, which make it difficult to investigate issues arising in adults, such as heterogeneity in symptoms and the timing of clinical manifestations. The All of Us Research Program is a nationwide precision medicine initiative that provides an important cohort consisting of 393,596 participants with both health records, survey data, and whole genome sequencing data, making it a unique resource to study ASD adults and other complex diseases such as cancer. In this study, we identified 1,049 ASD cases and 22,777 non-autistic controls from the All of Us cohort. Autistic adults presented a high rate (over 90%) of co-occurring psychiatric conditions, with over 60% receiving a delayed ASD diagnosis after other mental disorders. Comparison analysis of the 739 matched cases and controls showed statistically significant differences in various sociodemographic factors, including marriage, employment rates, and annual income.

## Introduction

Autism Spectrum Disorder (ASD) is a complex neurodevelopmental condition estimated to have a high heritability of about 83% [1]. According to the most recent US Autism and Developmental Disabilities Monitoring (ADDM) network from CDC, approximately 1 in 31 (3.2%) 8-year-old children were identified with ASD in 2022 [2]. However, patients with ASD can show heterogeneous clinical manifestations, with high variability in intellectual levels, language as well as communication abilities, and level of support needs for repetitive or restrictive behaviors [3]. Frequently, co-occurring conditions such as attention-deficit/hyperactivity disorder (ADHD), anxiety, epilepsy, and sleep problems further complicate the clinical picture [4,5]. Symptom manifestation also varies in timing, with some infants showing atypical behaviors by 6–12 months, others experiencing regression between 15–30 months, and other patients not being recognized until school age or adolescence [5–9]. Despite these well-documented heterogeneities, our understanding of the underlying biological and developmental mechanisms remains limited, highlighting the need for further studies to clarify how genetic and environmental factors influence different clinical trajectories.

The All of Us Research Program [10] offers a unique opportunity to address these challenges. This large, diverse, and rich cohort includes a substantial number of adults with ASD diagnoses, whole-genome sequencing (WGS) data, and extensive electronic health record (EHR) information, along with detailed environmental exposure data. While several large-scale ASD studies, including those from the Simons Simplex Collection (CSC) [11], MSSNG [12], and SPARK [13] cohorts, have mainly focused on early diagnosis, the All of Us cohort provides a special opportunity for us to explore late clinical manifestations of ASD and their potential interactions with biological and environmental factors.

This report examines ASD participants in the All of Us Research Program. We created an initial summary that highlights existing evidence gaps in adult-focused research and outlines the potential analytic approaches that could be applied to All of Us data to address these challenges. We also explore the availability of matched controls in the database, considering major confounding factors such as age, sex, race, and living environment. We will further investigate the potential genetic mechanisms by analyzing the whole genome sequencing data alongside clinical and demographic variables in future studies.

## Results

### Cohort selection flowchart

The All of Us Program is a comprehensive database that integrates various data sources, including EHRs, surveys, physical measurements, and genomics data across a large and diverse group of participants. Starting with the initial cohort of 633,537 participants in the All of Us v8 dataset, we selected a base cohort of 393,596 participants who had both EHRs and survey data available. (**Figure 1**)

**Figure 1.**
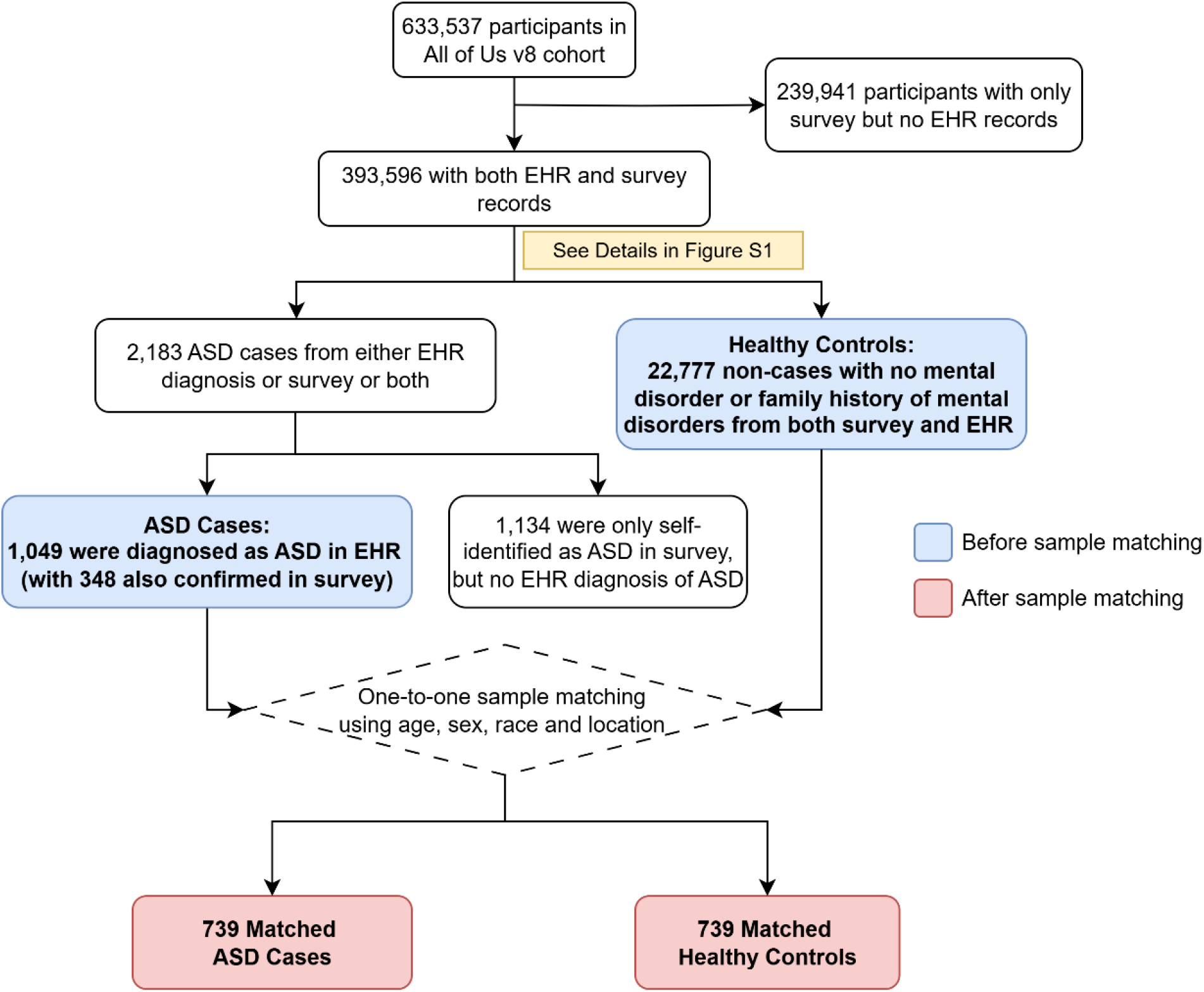
Study cohort setup flowchart. The flowchart illustrates the procedures to identify ASD cases and match them with healthy controls in the All of Us Research Program. The primary cohorts for matching are shown in blue shade and the one-to-one matched cohorts are shown in red shade. Detailed steps for obtaining the healthy controls can be found in **Supplementary Information Figure S1**.

Potential ASD cases included individuals with either an ASD diagnosis record from EHR or self-identification as having ASD in the “Personal and Family Health History” survey. However, we excluded 1,134 samples who were only self-identified but not clinically reported. This led to a final cohort of 1,049 cases with evidence of ASD or its underlying conditions (**SI Table S1**) confirmed through clinical diagnosis in EHRs. (**Figure 1**) Neurotypical controls totaling 22,777 participants were identified through careful screening steps (**SI Figure S1**) to select individuals who reported no personal or family history of ASD or other psychiatric disorders from either EHRs or surveys.

### Age patterns of clinical diagnosis and psychiatric comorbidity in ASD cases

Our investigation of 1,049 ASD cases showed that 85.03% of individuals were first diagnosed with ASD in adulthood, with a median age of 29 years and a wide age range from as young as 3 years old to the late 70s (**Table 1**). More than half (56.43%) of the initial diagnoses occurred between ages 18 and 39, nearly 17% at age 50 or older, and 6.39% at age 60 or older. The median time between their first ASD diagnosis and survey enrollment is 2 years, with over 80% of participants enrolled after their diagnosis.

**Table 1.** Personal and family ASD history and psychiatric comorbidity patterns in 1,049 ASD cases. This table summarizes patterns of ASD diagnosis and non-ASD psychiatric conditions in EHRs within the selected case cohort of 1,049 people. To comply with the All of Us policy, participant counts that are less than 20 are reported as “<20”, and the second lowest number, if higher than 20, is also masked with a lower bound to avoid the masked count values being derived.

| Variables | All cases (%)<br>N=1,049 |
| --- | --- |
| **Age at first diagnosis of ASD**<br>**(from EHR records)** |  |
| Mean ± SD | 32.46 ± 15.45 |
| Median (Min, Max) | 29 (3, 76) |
| 0-9 | >21 (>2.00%) |
| 10-17 | 133 (12.68%) |
| 18-29 | 374 (35.65%) |
| 30-39 | 218 (20.78%) |
| 40-49 | 123 (11.73%) |
| 50-59 | 110 (10.49%) |
| 60-69 | 50 (4.77%) |
| Over 70 | <20 (<1.91%) |
| **Years between survey and the first**<br>**diagnosis of ASD** |  |
| Mean ± SD | 3.67 ± 5.65 |
| Median (Min, Max) | 2 (-5, 34) |
| <0 | 202 (19.26%) |
| [0, 5) | 477 (45.47%) |
| [5, 10) | 224 (21.35%) |
| 10+ | 146 (13.92%) |
| **Concurrent other mental disorders** |  |
| Yes | 970 (92.47%) |
| No | 79 (7.53%) |
| **Diagnosis order** |  |
| ASD diagnosed first | 310 (29.55%) |
| Other mental disorders diagnosed first | 660 (62.92%) |
| Only ASD, no other mental disorders | 79 (7.53%) |
| **Years between ASD diagnosis and**<br>**other mental disorders** |  |
| Mean ± SD | 6.05 ± 5.38 |
| Median (Min, Max) | 5 (1, 39) |
| <0 | 165 (15.73%) |
| [0, 5) | 596 (56.82%) |
| [5, 10) | 186 (17.73%) |
| 10+ | 83 (7.91%) |
| **First-degree family history of ASD** |  |
| Yes | 118 (11.25%) |
| No | 250 (23.83%) |
| Missing | 681 (64.92%) |

Our results also showed a high prevalence of psychiatric comorbidity in ASD cases. 92.47% of the cohort had more than one mental disorder conditions diagnosed besides ASD, while only 79 (7.53%) people had ASD as their sole mental condition. In addition, the majority (62.92%) of the ASD diagnosis occurred after other psychiatric conditions had been diagnosed, with a time lag average of about 6 years. (**Table 1**) In terms of the initial diagnosis age of ASD, patients without comorbidity (median diagnosis age: 28 years) were significantly older than comorbid patients with ASD diagnosed first (median diagnosis age: 23 years; P = 0.0094), but significantly younger than comorbid patients with other mental conditions diagnosed first (median diagnosis age: 32 years; P = 0.0356). (**Figure 2**)

**Figure 2.**
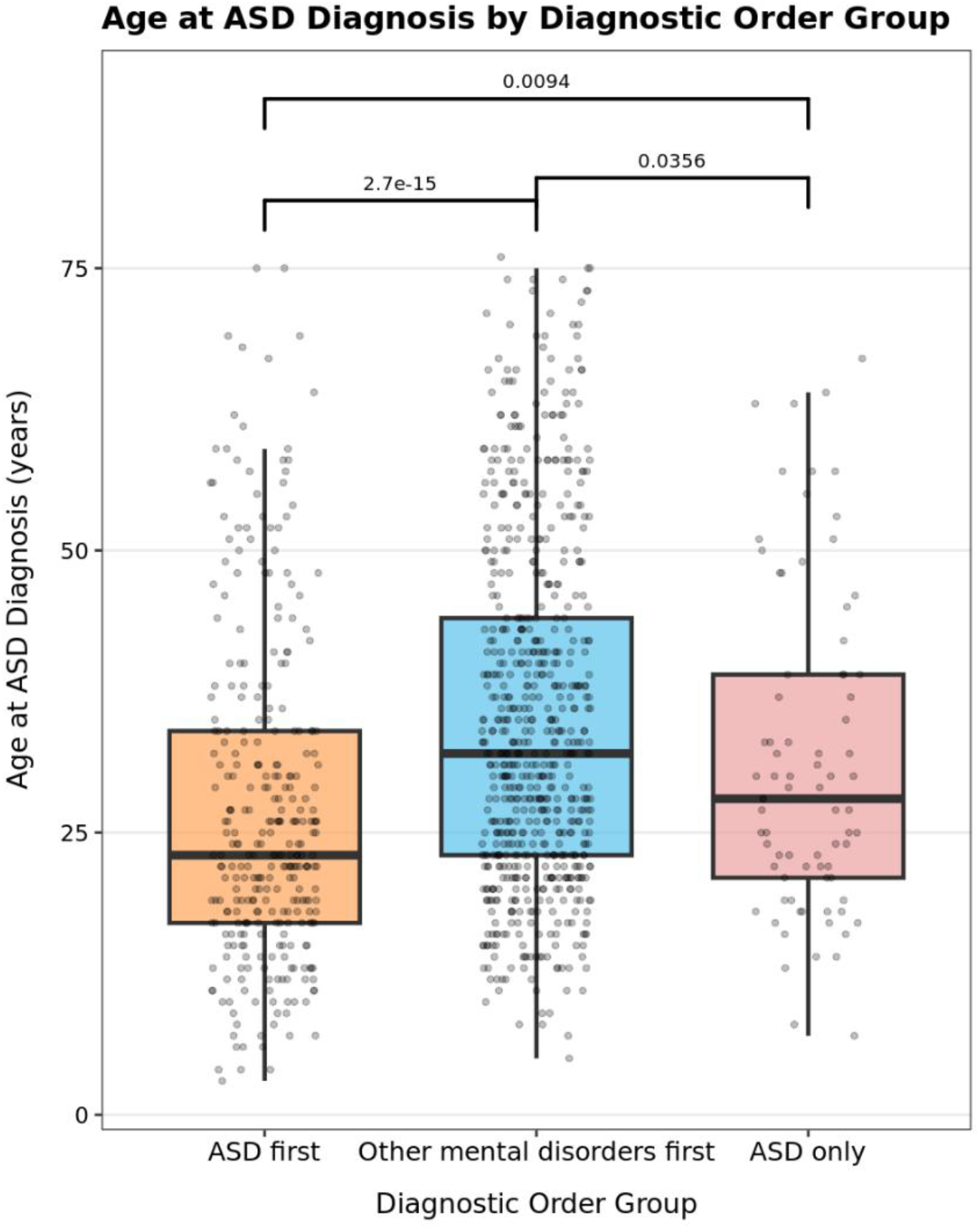
Distribution of age at ASD diagnosis by Diagnosis order. This boxplot compares age at first ASD diagnosis across three diagnosis order groups: ASD diagnosed before other mental disorders (“ASD first”), other mental disorders diagnosed before ASD (“Other mental disorders first”), or ASD only with no other mental disorders (“ASD only”). (**Table 1**) Each dot represents age of a participant. Wilcoxon tests were performed for pairwise comparisons across the three groups, with their p-values labeled above the brackets.

By comparing the age distribution of ASD diagnosis between males and females using the 739 matched ASD cases (with 367 female and 372 males), the Kaplan-Meier curves revealed a clear difference between the two sex groups. Males tend to be diagnosed with ASD at a younger age, with 17.74% of the males diagnosed before age of 20 compared to 15% in females. However, the diagnosis rate grows more rapidly for females, especially during age of 30-50, with 37.87% females diagnosed during this period compared to 31.18% for males. (**Figure 3**) A log-rank test to compare the difference had a p-value of 0.023, indicating significant difference in the diagnosis age between sex.

**Figure 3.**
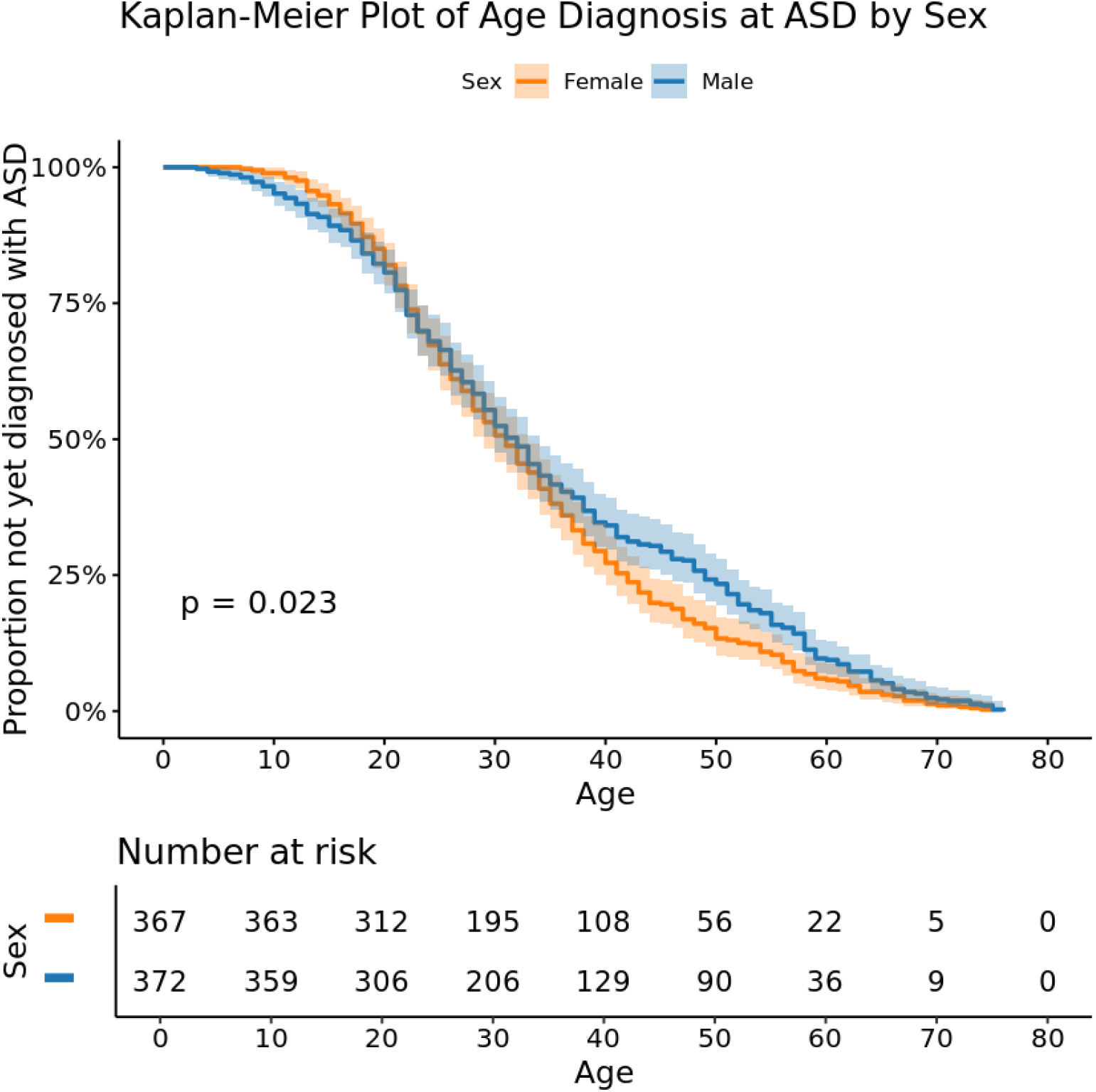
Kaplan-Meier curve of age at ASD diagnosis by sex. The Kaplan-Meier curve showed the aggression to ASD along age by female and male separately. Shown in the x-axis and y-axis are the age at first ASD diagnosis and the survival probability of no diagnosis of ASD, respectively. Only 739 matched ASD cases with binary sex (Female/Male) were used for this plot. The shaded areas around the curve are the 95% confidence interval. The p-value of 0.023 came from the log-rank test between female and male.

### Demographic and sociodemographic characteristics of analytical cohorts

Among all 1,049 ASD cases and 22,777 neurotypical controls, the mean age of participants in the control group was 57.7 years (±17.1), which was significantly higher than the mean age in the case group of 36.1 years (±14.7). Control samples tend to be older, with over 50% of controls over 60 years old, compared to only about 11% in the case group. For gender, there were 514 (49.00%) females and 523 (49.86%) males among cases, while among controls, there were 11,601(50.93%) males and 11,034 (48.44%) females. Regarding race, 734 cases (69.97%) were White, compared to 13,098 controls (57.51%).

We then applied one-to-one sample matching based on age at survey, sex at birth, race and physical location at the state level, resulting in balanced case-control sample sets for comparison analysis. This resulted in 739 ASD cases and 739 healthy neurotypical controls, totaling 1,478 samples in the final analytical cohort (**Figure 1**).

After matching, the mean age in the case and control groups was 37.9 years (±15.0) and 38.0 years (±14.9), respectively. Race, state, and sex were exactly matched between the two groups. 367 pairs (49.66%) were female, and 372 pairs (50.34%) were male. In terms of race, 656 pairs (88.77%) were White, and 69 pairs (9.34%) were Black. Geographically, 158 pairs (21.38%) were from the West, 204 pairs (27.60%) from the Midwest, 282 pairs (38.16%) from the Northeast, and 95 pairs (12.86%) from the South. [14] More details are presented in **Table 2**.

**Table 2.** Demographic characteristics of ASD cases and healthy controls before and after samples matching. This table summarizes the four demographic variables that were used for sample matching between ASD cases and healthy controls. The geographic regions of the participants’ location were determined using the CDC standards and were derived from the exact zip codes [19]. To comply with the All of Us policy, participant counts that are less than 20 are reported as “<20”, and the second lowest number, if higher than 20, is also masked with a lower bound to avoid the masked count values being derived.

| Variables | Before sample matching |  | After sample matching |  |
| --- | --- | --- | --- | --- |
|  | All cases (%)<br>N=1,049 | All controls (%)<br>N=22,777 | Matched cases (%)<br>N=739 | Matched controls (%)<br>N=739 |
| **Age at enrollment** |  |  |  |  |
| Mean ± SD | 36.13 ± 14.73 | 57.70 ± 17.069 | 37.87 ± 14.99 | 38.04 ± 14.85 |
| Median (Min, Max) | 32 (18, 80) | 61 (18, 104) | 33 (18, 80) | 33 (19, 83) |
| 18-29 | 451 (42.99%) | 1,982 (8.7%) | 272 (36.81%) | 260 (35.18%) |
| 30-39 | 261 (24.88%) | 2,516 (11.05%) | 201 (27.20%) | 214 (28.96%) |
| 40-49 | 129 (12.30%) | 2,558 (11.23%) | 95 (12.86%) | 95 (12.86%) |
| 50-59 | 94 (8.96%) | 3,670 (16.11%) | 81 (10.96%) | 80 (10.83%) |
| 60-69 | 74 (7.05%) | 5,762 (25.30%) | 57 (7.71%) | 57 (7.71%) |
| Over 70 | 40 (3.81%) | 6,289 (27.61%) | 33 (4.47%) | 33 (4.47%) |
| **Sex at birth** |  |  |  |  |
| Female | 514 (49.00%) | 11,601 (50.93%) | 367 (49.66%) | 367 (49.66%) |
| Male | 523 (49.86%) | 11,034 (48.44%) | 372 (50.34%) | 372 (50.34%) |
| Other/Intersex | <20 (<1.91%) | <20 (<0.09%) | 0 | 0 |
| Missing | <20 (<1.91%) | >122 (>0.54%) | 0 | 0 |
| **Race** |  |  |  |  |
| White | 734 (69.97%) | 13,098 (57.51%) | 656 (88.77%) | 656 (88.77%) |
| Black | 83 (7.91%) | 3,024 (13.28%) | 69 (9.34%) | 69 (9.34%) |
| Asian | <20 (<1.91%) | 1,294 (5.68%) | <20 (<2.71%) | <20 (<0.09%) |
| American Indian or Alaska Native | <20 (<1.91%) | 223 (0.98%) | <20 (<2.71%) | <20 (<0.09%) |
| Middle Eastern or North African | <20 (<1.91%) | 181 (0.79%) | 0 | 0 |
| Other/Mixed | 109 (10.39%) | 836 (3.67%) | 0 | 0 |
| Missing | 90 (8.58%) | 4,121 (18.09%) | 0 | 0 |
| **Locations** |  |  |  |  |
| West | 223 (21.26%) | 7,632 (33.51%) | 158 (21.38%) | 158 (21.38%) |
| Midwest | 243 (23.16%) | 6,431 (28.23%) | 204 (27.60%) | 204 (27.60%) |
| Northeast | 370 (35.27%) | 4,004 (17.58%) | 282 (38.16%) | 282 (38.16%) |
| South | 149 (14.20%) | 4,171 (18.31%) | 95 (12.86%) | 95 (12.86%) |
| Missing | 64 (6.10%) | 539 (2.37%) | 0 | 0 |

### Personal health characteristics of analytical cohorts

Table 3 summarizes all variables with the matched samples. Among these samples, 499 (67.52%) of control samples were classified as “Healthy Weight” (18.5 ≤ BMI < 25) or “Overweight” (25 ≤ BMI < 30) according to CDC categories, while most cases (548 cases (74.15%)) fell into the “Overweight” (25 ≤ BMI <30) or “Obesity” groups (BMI ≥ 30).

Regarding education, 657 individuals (88.90%) in the control group attended college, compared to 499 patients with ASD (67.52%). Employment and household income also differed between groups. Among controls, 568 controls (76.86%) were still employed compared to 308 in cases (41.68%). Controls generally reported higher income, with 517 (69.96%) earning more than $25,000 annually compared to 303 in cases (41.00%). Moreover, 98 controls (13.26%) had income exceeding $200,000, while only 26 cases (3.52%) reported income above this level. 389 controls (52.64%) were married or lived with partners, whereas only 207 cases (28.01%) married or lived with partners. 298 controls (40.32%) never married, while 429 cases (58.05%) never married.

**Table 3.** Sociodemographic and personal health characteristics of matched cases and controls. This table summarizes the distribution of sociodemographic and personal health factors among matched cases and controls. Paired t-tests and *χ*^2^ tests were performed to compare differences between the two groups. (*: the statistically significant P values with threshold of 0.01 are shown in bold.) To comply with the All of Us policy, participant counts that are less than 20 are reported as “<20”, and the second lowest number, if higher than 20, is also masked with a lower bound to avoid the masked count values being derived.

| Variables | After sample matching |  | P value* |
| --- | --- | --- | --- |
|  | Matched cases (%)<br>N=739 | Matched controls (%)<br>N=739 |  |
| **BMI (kg/m<sup>2</sup>)** |  |  |  |
| Mean ± SD | 31.89 ± 9.20 | 27.73 ± 6.94 | **$7.19 \times 10^{-22*}$** |
| Median (Min, Max) | 30.28 (16.36, 71.31) | 26.34 (14.18, 76.99) |  |
| <18.5 | <20 (<2.71%) | <20 (<2.71%) |  |
| [18.5, 25) | 161 (21.79%) | 278 (37.62%) |  |
| [25, 30) | 186 (25.17%) | 221 (29.91%) |  |
| [30, 40) | 231 (31.26%) | 160 (21.65%) |  |
| $\geq 40$ | 131 (17.73%) | >22 (>2.98%) | |
| Missing | <20 (<2.71%) | 38 (5.14%) |  |
| **Ethnicity** |  |  |  |
| Hispanic | 30 (4.06%) | 49 (6.63%) | $3.74 \times 10^{-2}$ |
| Non-Hispanic | 709 (95.94%) | 690 (93.37%) |  |
| **Country of birth** |  |  |  |
| USA | 713 (96.48%) | 664 (89.85%) | **$1.62 \times 10^{-7*}$** |
| Other | >6 (>0.81%) | >55 (>7.44%) |  |
| Missing | <20 (<2.71%) | <20 (<2.71%) |  |
| **Highest level of education** |  |  |  |
| Advanced degree | >97 (>13.13%) | 248 (33.56%) | **$3.80 \times 10^{-34*}$** |
| College graduate | 176 (23.82%) | 253 (34.24%) |  |
| College one to three | 219 (29.63%) | 156 (21.11%) |  |
| High school or lower | 227 (30.72%) | >62 (>8.39%) |  |
| Missing | <20 (<2.71%) | <20 (<2.71%) |  |
| **Language other than English** |  |  |  |
| Yes | 34 (4.60%) | 66 (8.93%) | $2.94 \times 10^{-1}$ |
| No | 318 (43.03%) | 477 (64.55%) |  |
| Missing | 387 (52.37%) | 196 (26.52%) |  |
| **Annual household income** |  |  |  |
| $\geq 200k$ | 26 (3.52%) | 98 (13.26%) | **$2.73 \times 10^{-43*}$** |
| [75k, 200k) | 105 (14.21%) | 223 (30.18%) |  |
| [25k, 75k) | 172 (23.27%) | 196 (26.52%) |  |
| < 25k | 295 (39.92%) | 84 (11.37%) |  |
| Missing | 141 (19.08%) | 138 (18.67%) |  |
| **Employment status** |  |  |  |
| Employed | 308 (41.68%) | 568 (76.86%) | **$1.55 \times 10^{-39*}$** |
| Not employed | 397 (53.72%) | >151 (>20.43%) |  |
| Missing | 34 (4.60%) | <20 (<2.71%) |  |
| **Marital status** |  |  |  |
| Married or with partner | 207 (28.01%) | 389 (52.64%) | $7.27 \times 10^{-43*}$ |
| Never married | 429 (58.05%) | 298 (40.32%) |  |
| Divorced or separated | 76 (10.28%) | 39 (5.28%) |  |
| Widowed | <20 (<2.71%) | <20 (<2.71%) |  |
| Missing | <20 (<2.71%) | <20 (<2.71%) |  |
| **Cigarette** |  |  |  |
| Never | 538 (72.80%) | 611 (82.68%) | $1.96 \times 10^{-6*}$ |
| Current | 81 (10.96%) | >19 (>2.57%) |  |
| Former | 99 (13.40%) | 89 (12.04%) |  |
| Missing | 21 (2.84%) | <20 (<2.71%) |  |
| **Vaping** |  |  |  |
| Never | 541 (73.21%) | 581 (78.62%) | $5.83 \times 10^{-2}$ |
| Current | 44 (5.95%) | >16 (>2.17%) |  |
| Former | 131 (17.73%) | 122 (16.51%) |  |
| Missing | 23 (3.11%) | <20 (<2.71%) |  |
| **Alcohol** |  |  |  |
| Never | 128 (17.32%) | 67 (9.07%) | $8.08 \times 10^{-20*}$ |
| Current | 153 (20.70%) | >48 (>6.50%) |  |
| Former | 434 (58.73%) | 604 (81.73%) |  |
| Missing | 24 (3.25%) | <20 (<2.71%) |  |
| **General health** |  |  |  |
| Excellent | >45 (>6.09%) | 117 (15.83%) | $9.26 \times 10^{-54*}$ |
| Very good | 135 (18.27%) | 368 (49.80%) |  |
| Good or fair | 472 (63.87%) | 243 (32.88%) |  |
| Poor | 67 (9.07%) | <20 (<2.71%) |  |
| Missing | <20 (<2.71%) | <20 (<2.71%) |  |
| **General physical health** |  |  |  |
| Excellent | >34 (>4.60%) | 108 (14.61%) | $8.00 \times 10^{-46*}$ |
| Very good | 137 (18.54%) | 332 (44.93%) |  |
| Good or fair | 467 (63.19%) | 280 (37.89%) |  |
| Poor | 81 (10.96%) | <20 (<2.71%) |  |
| Missing | <20 (<2.71%) | <20 (<2.71%) |  |
| **General mental health** |  |  |  |
| Excellent | >36 (>4.87%) | 190 (25.71%) | $3.12 \times 10^{-81*}$ |
| Very good | 120 (16.24%) | 332 (44.93%) |  |
| Good or fair | 468 (63.33%) | 215 (29.09%) |  |
| Poor | 95 (12.86%) | <20 (<2.71%) |  |
| Missing | <20 (<2.71%) | <20 (<2.71%) |  |
| **General quality of life** |  |  |  |
| Excellent | 64 (8.66%) | 214 (28.96%) | $1.97 \times 10^{-6*}$ |
| Very good | 168 (22.73%) | 359 (48.58%) |  |
| Good or fair | 443 (59.95%) | 163 (22.06%) |  |
| Poor | >44 (>5.95%) | <20 (<2.71%) |  |
| Missing | <20 (<2.71%) | <20 (<2.71%) |  |
| **Feel doctor or nurse not listening during appointment** |  |  |  |
| Never | 68 (9.20%) | 277 (37.48%) | $1.04 \times 10^{-29*}$ |
| Rarely or sometimes | 206 (27.88%) | 247 (33.42%) |  |
| Always or most of the time | 78 (10.55%) | <20 (<2.71%) |  |
| Missing | 387 (52.37%) | >195 (>26.39%) |  |

In the items about overall health (“general health,” “general physical health,” “general mental health,” and “general quality of life”), almost all participants in the control group rated themselves as “fair” or better, with more than 60% reporting “very good” or “excellent”. In contrast, only 25% to 30% of case participants reported “very good” or “excellent”, around 60% felt “fair” or “good”, and approximately 10% of cases reported “poor” health or quality of life.

### Comparison analysis of characteristics

Among 16 sociodemographic variables and personal health characteristics, our results showed statistically significant differences between ASD patients and controls in 13 variables, with insignificant difference in “ethnicity,” “language other than English,” and “vaping”.

Patients with ASD had significantly higher BMI (P = 7.19 × 10^-22^), significantly different distributions of household income (P = 2.73 × 10^-43^) and advanced education (P = 3.80 × 10^-34^) than controls. The percentage of employment also differed remarkedly (P = 1.55 × 10^-39^) among individuals with ASD (41.68%) compared to the controls (76.86%). Marital status also differed markedly (P = 7.27 × 10^-43^) between cases and controls. Differences were further observed in health-related measures with very small p values, including “general health”, “physical health,” and “mental health.” Not surprisingly, the quality of life and patient–provider communication was rated more favorably in the control group compared to ASD cases. Additionally, significant differences were also present for “country of birth” (P = 1.62 × 10^-7^), “cigarette use” (P = 1.96 × 10^-6^) and “alcohol use” (P = 8.08 × 10^-20^). Please see **Table 3** for the detailed results.

## Discussion

Since ASD affects neurodevelopment early in life, early diagnosis and intervention are important and desired. Thus, most cohort studies have focused on children, with far fewer studies on adults. Primarily a nationwide precision medicine initiative, the All of Us Research Program has collected diverse health data, including EHR and survey responses, to examine how genetic, environmental, and lifestyle factors shape health and disease, such as cancer. With 633,540 adult participants and 3,558 individuals with ASD enrolled, it provides an excellent resource for studying autism in adults. Within the program, an important cohort consists of 393,596 participants with both EHR and survey data, including 2,183 ASD cases, with 1,049 diagnosed in EHR and 1,134 self-identified.

Among individuals diagnosed with ASD in EHR, 49.38% of their initial diagnoses occurred at age 30 or older. Such a tendency for late ASD diagnosis in adulthood may reflect challenges in timely detection and greater awareness about ASD among clinical providers. We further identified 739 matched pairs of ASD patients and non-autistic controls and explored their differences, along with the comparison of basic and sociodemographic characteristics. Notably, patients with ASD had statistically significant lower rates of marriage and employment, with much lower annual income. While these findings are consistent with the literature, it is interesting to observe that there were higher rates of cigarette smoking and alcohol use in the ASD case population, although the difference in vaping was not statistically significant.

Our results contribute to the growing body of literature on autism, highlight the importance of sociodemographic and personal health characteristics in ASD patients, and inform future research and interventions for autistic individuals. However, they also post a few challenges to our future studies. The *All of Us* Research Program established a nationwide community of participants primarily through health provider organizations, with regions that have strong clinical networks, such as California and Arizona, contributing disproportionately large numbers of samples. In the Basics survey, more than 50% of participants self-identified as White. Consequently, our study cohort is skewed towards White participants (58.1%) and residents in Arizona and California (28.5%). These imbalances may limit representation of minority ASD populations. Some reports [15,16] suggest that minorities may have a higher prevalence of ASD than White populations. Future analyses should therefore prioritize larger, more geographically and demographically diverse samples to improve representation.

ASD is frequently accompanied by one or more psychiatric comorbidities[17], with 70% of ASD youth having at least one co-occurring disorder and 41% having two or more[18]. In the cohort from All of Us, the prevalence of psychiatric comorbidity is even higher, with over 92% of ASD adults having at least one co-occurring psychiatric disorder and 85.51% presenting multiple psychiatric comorbidities. Common co-occurring conditions shown in All of Us include Anxiety Disorders (57.96%), Depressive Disorders (46.04%), Attention-Deficit/Hyperactivity Disorder (ADHD; 39.66%), Obsessive-Compulsive Disorders (OCD; 12.01%), and Intellectual Disability (ID; 5.91%). While such high rate of comorbidities in ASD adults provides an opportunity to study the interplay between ASD and other psychiatric disorders, it also poses analytic challenges that require carefully designed models and methodologies. For instance, identifying genetic risk factors for ASD may require innovative models that account for both those shared with other mental disorders and those unique to ASD. The fact that ASD is typically preceded by mental health difficulties with an average lag of six years adds further complexity to modeling.

With 600 cases and 635 controls (out of 739 in each group) having the whole genome sequencing data, the All of Us dataset offers a unique opportunity to identify the genetic variants associated with ASD, which can help us estimate the polygenic risk score (PRS) for risk stratification, explore gene-environment interactions, facilitate more accurate diagnosis and personalized treatment strategies. In addition, longitudinal information can be used to model the patients’ health trajectories, identify protective factors, and evaluate the treatment outcomes.

## Material and Methods

### Cohort Selection

The study cohorts were selected from the All of Us Research Program participants who contributed both electronic health records (EHR) and matched survey responses across six survey types, including the core surveys (“The Basics,” “Lifestyle,” “Overall Health”) and follow-up surveys (“Personal and Family Health History,” “Healthcare Access and Utilization,” and “Social Determinants of Health”). The selection process involved multiple screening steps with clearly defined criteria for ASD cases and non-autistic controls (**Figure 1**).

Classification of ASD cases requires either a documented diagnosis of ASD or related conditions from the EHR data. Specifically, participants were considered as cases if they had been diagnosed with ASD under *SNOMED code 35919005* (or *OMOP concept ID 439776*) and had completed at least those three core surveys. A detailed breakdown of the underlying conditions of *SNOMED 35919005* can be found in **Supplementary Information (SI Table S1)**. Since multiple studies have shown that ASD often co-occurs with other psychiatric conditions [19–22], we defined non-autistic controls as those who, as well as their first-degree family members, showed no evidence of autism or other mental disorders, verified through EHRs and the “Personal and Family Health History” survey. The selected cases and controls were combined for descriptive analyses of demographic, sociodemographic, and personal health characteristics, providing an initial overview of the study cohort.

### Data and variable extraction

Some participant information was extracted from EHR and survey data, including “sex at birth,” “race,” and “ethnicity.” “Age at survey” was calculated based on the date of birth and the first enrollment date in “The Basic” survey. The details of these variables are presented in **SI Table S3**.

BMIs, heights, and weights were extracted from the in-lab physical measurement records. The records were merged by individual and measurement time, keeping only those with either provided BMI values or time-matched height and weight measurements. Measuring units of all measures were unified before data cleaning. To reduce the impact of extreme values, we scanned the records of BMI, height, and weight in individual-month levels, and used 6 times of the mean absolute deviation (MAD) to identify potential outliers. Implausible BMI values that are less than 12 or greater than 100 were treated as missing. For participants with both height and weight, we recalculated BMI and checked its consistency with the reported value. For each person, BMI values were averaged within each year, and the annual average BMI closest to the survey age (within 3 years) was selected. Self-reported BMI was used only if clinical data were unavailable.

Besides age, sex, race, ethnicity, and BMI, we also extracted additional information from the survey data. From “The Basic” survey, we collected data from questionnaires including “the highest level of education,” “country of birth,” “annual household income,” “marital status,” and “employment status.” The responses to questions like “feel doctor or nurse not listening during appointment” and “language other than English spoken at home” were gathered from the “Social Factors of Health” survey. From the “Overall Health” survey, we selected measures such as “general quality of life,” “general mental health,” “general physical health,” and “general health”. Information on “Cigarettes use,” “vaping,” and “alcohol consumption” was obtained from the “Lifestyle” survey. Details of these questionnaires and their designs are presented in **SI Table S4**.

### Data preprocessing

After extracting variables from the dataset, we excluded participants with missing values in race, sex at birth, and physical location (at the state level) in both cases and control groups, resulting in 17,506 controls and 826 cases. For other variables, responses such as “Skip,” “Prefer not to answer,” or other non-informative categories were treated as missing values.

### Sample matching

We performed one-to-one sample matching using four demographic variables (“age at survey enrollment,” “sex at birth,” “race,” and “locations at the state level”) to ensure a balanced comparison between patients with ASD and neurotypical controls. Participants with ambiguous categories or missing values in age and sex were excluded before matching. Exact matches were required for sex, race, and state. For age, we used a greedy nearest neighbor matching approach without replacement with dynamic caliper widths, which have been shown to produce higher propensity scores between matched samples [23]. Specifically, we began with exact matching for age, then applied a progressively increasing caliper width of one year to the remaining unmatched samples. The maximum caliper width was capped at ±3 years to maintain close age matching between groups. Participants that could not be matched within this rule were omitted from subsequent statistical analyses. While exact age matching resulted in 568 pairs, increasing the caliper width to 1, 2, and 3 years yielded 671, 714, and 739 pairs, respectively. We summarized the demographic characteristics of cases and controls before and after sample matching, with an age caliper width of 3 years, in **Table 1**.

### Statistical Analysis

After matching, we compared cases and controls across different sociodemographic and health characteristics and performed paired t tests on continuous variables (i.e., BMI) and *χ*^2^ tests for other categorical variables (**Table 2**). Specifically for cases, we performed pairwise comparisons using Wilcoxon rank-sum tests to compare diagnosis age distribution across three diagnostic sequential order groups− ASD first, other mental disorders first, ASD only (with no other mental conditions). We generated Kaplan-Meier survival curves to compare the diagnosis age of ASD between male and female, with a log-rank test to evaluate the significance of the difference.

## Data Availability

All data produced in the present study are available upon request to the the National Institutes of Health All of Us Research Program.

## Acknowledgements

We gratefully acknowledge All of Us participants for their contributions, without whom this research would not have been possible. We also thank the National Institutes of Health’s All of Us Research Program for making available the participant data examined in this study.

Research reported in this publication was supported in part by the National Institute on Aging under award number R01AG080917 and R01AG080917-02S1, the National Cancer Institute of the National Institutes of Health under award number P30CA062203 and the UC Irvine Comprehensive Cancer Center using UCI Anti-Cancer Challenge funds. The content is solely the responsibility of the authors and does not necessarily represent the official views of the National Institutes of Health or the Chao Family Comprehensive Cancer Center.

