## Supplemental Materials for "Characterizing Autism Spectrum Disorder in the All of Us Research Program"

### Supplementary Information

#### Figure S1. Flowchart of omitted part in Figure 1

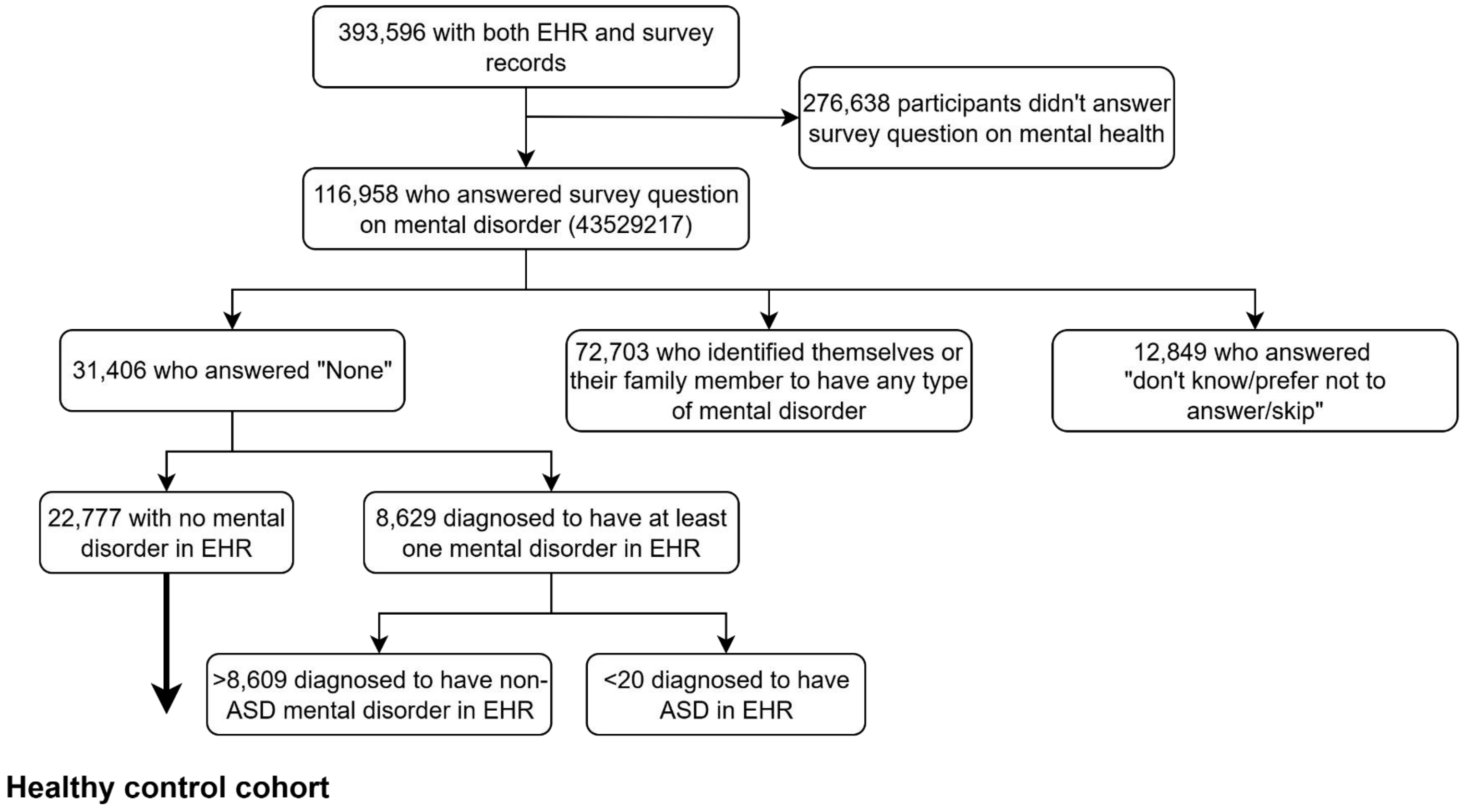

**Table S1. Hierarchical breakdown of ASD and its related condition in All of Us**

Autism spectrum disorder (*SNOMED 35919005*)

- Asperger's disorder (*SNOMED 23560001*)
- Autistic disorder (*SNOMED 408856003*)
    - High-functioning autism (*SNOMED 702732007*)
    - Infantile autism (*SNOMED 408857007*)
        - Active infantile autism (*SNOMED 191689008*)
        - Residual infantile autism (*SNOMED 191690004*)
- Childhood disintegrative disorder (*SNOMED 71961003*)
- Pervasive developmental disorder of residual state (*SNOMED 39951000119105*)
    - Residual infantile autism (*SNOMED 191690004*)
- Rett's disorder (*SNOMED 68618008*)

**Table S2. Counts of different mental disorders in the All of Us Research Program**

| Name | SNOMED | Roll-up Count |
|---|---|---|
| Mental disorder | 74732009 | 177,841 |
| Chronic mental disorder | 128293007 | 48,745 |
| Mental disorder due to drug | 442351006 | 43,553 |
| Organic mental disorder | 111479008 | 39,628 |
| Psychoactive substance-induced organic mental disorder | 11387009 | 25,510 |
| Developmental mental disorder | 129104009 | 17,091 |
| Neurosis | 111475002 | 11,484 |
| Cannabis-induced organic mental disorder | 77355000 | 10,716 |
| Opioid-induced organic mental disorder | 14784000 | 8,483 |
| Alcohol-induced organic mental disorder | 29212009 | 4,804 |
| Cocaine-induced organic mental disorder | 46975003 | 3,953 |
| Insomnia disorder related to another mental disorder | 24121004 | 2,250 |
| Mental disorders during pregnancy, childbirth and the puerperium | 199257008 | 2,171 |
| Nicotine-induced organic mental disorder | 30310000 | 1,281 |
| Intellectual disability | 110359009 | 1,149 |
| Mental disorder during pregnancy - baby delivered | 199259006 | 0 |
| Social problem not due to a mental disorder | 56098000 | 732 |
| Mental disorder in childhood | 3914008 | 497 |
| Mild intellectual disability | 86765009 | 0 |
| Transient organic mental disorder | 428703001 | 0 |
| Mental disorder in infancy | 24125008 | 286 |
| Social maladjustment | 67564005 | 0 |
| Pregnancy with mental disorders | 267320004 | 0 |
| Hallucinogen-induced organic mental disorder | 53050002 | 176 |
| Borderline intellectual disability | 77287004 | 0 |
| Moderate intellectual disability | 61152003 | 0 |
| Hypersomnia disorder related to another mental disorder | 89415002 | 0 |
| Chronic organic mental disorder | 425919003 | 66 |
| Specific nonpsychotic mental disorders following organic brain damage | 192069009 | 0 |
| Mental disorder AND/OR culture bound syndrome | 106015009 | 49 |
| Mental disorder in the puerperium - baby delivered | 199260001 | 0 |
| Mental disorder during pregnancy - baby not yet delivered | 199261002 | 0 |
| Mental disorder in adolescence | 26453000 | 28 |
| Inhalant-induced organic mental disorder | 61104008 | 20 |

| | | |
|---|---|---|
| Mental disorder in mother complicating pregnancy | 702711004 | <20 |
| Severe intellectual disability | 40700009 | 0 |
| Mental disorder in mother complicating childbirth | 10811201000119102 | <20 |
| Profound intellectual disability | 31216003 | 0 |
| Amphetamine-induced organic mental disorder | 83367009 | <20 |
| Mental disorder caused by methamphetamine | 12398361000119105 | <20 |
| Caffeine-induced organic mental disorder | 309279000 | <20 |

**Table S3. Details of personal information**

| Variables | Categories | Original answer |
|---|---|---|
| Sex at birth | Male | Male |
| | Female | Female |
| | Other | Intersex, No matching concept and None of These |
| | Missing | Prefer Not to Answer, Skip |
| Race | American Indian or Alaska Native | American Indian or Alaska Native |
| | Asian | Asian |
| | Black or African American | Black or African American |
| | Middle Eastern or North African | Middle Eastern or North African |
| | White | White |
| | Other | Native Hawaiian or Other Pacific Islander, More than One Population, None of These |
| | Missing | Prefer Not to Answer, None Indicated and Skip |
| Ethnicity | Hispanic | Hispanic or Latino |
| | Not Hispanic | Not Hispanic or Latino |
| | Other | Race Ethnicity None of These |
| | Missing | Prefer Not to Answer, Skip, |
| Age at survey | Continuous | Date of survey enrollment (the first-time enrollment the basic survey)-date of birth |
| BMI | Continuous | Derived from EHR data and matched with Date of survey enrollment |

**Table S4. Details of survey information**

| Variables | Survey Source | Question (question id) | Values | Original answer (answer id) |
|---|---|---|---|---|
| Highest Level of Education | The Basics | What is the highest grade or year of school you completed? (1585940) | Advanced Degree | Advanced Degree (1585948) |
| | | | College Graduate | College Graduate (1585947) |
| | | | College One to Three | College One to Three (1585946) |
| | | | High School or Lower | One Through Four (1585942), Five through Eight (1585943), Nine through Eleven (1585944), Twelve or GED (1585944) Never Attended (1585941) |
| | | | Missing | Skip (903096), Prefer Not to Answer (903079), No Response |
| Country of Birth | The Basics | In what country were you born? (1586135) | USA | USA (1586136) |
| | | | Other | Other (903070) |
| | | | Missing | Skip (903096), No response |
| Annual household income | The Basics | What is your annual household income from all sources? (1585375) | More than 200K | More 200k (1585384) |
| | | | 75K-200K | 75k~100k (1585381), 100k~150k (1585382) 150k~200k (1585383) |
| | | | 25K-75K | 25k~35k (1585378), 35k~50k (1585379) 50k~75k (1585380) |
| | | | Less than 25K | less 10k (1585376), 10k~25k (1585377) |
| | | | Missing | Skip (903096), Prefer Not to Answer (903079), No Response |
| Marital | The Basics | What is your current marital status? (1585892) | Married or with Partner | Married (1585893), Living with Partner (1585898) |
| | | | Never Married | Never Married (1585897) |
| | | | Divorced or Separated | Divorced (1585894), Separated (1585896) |
| | | | Widowed | Widowed (1585895) |
| | | | Missing | Skip (903096), Prefer Not to Answer (903079), No Response |

| | | | | |
|---|---|---|---|---|
| Employment status | The Basics | What is your current employment status? Please select 1 or more of these categories. (1585952) | Employed | If choices include: Employed for Wages (1585953), Self Employed (1585954) |
| | | | Unemployed | If none above and choices include: Out of Work One or More (1585955), Out of Work Less Than One (1585956), Homemaker (1585957), Student (1585958), Retired (1585959), Unable to Work (1585960) |
| | | | Missing | Skip (903096), Prefer Not to Answer (903079), No Response |
| Feel Doctor or Nurse Not Listening During Appointment | Social Factors of Health | How often do you feel like a doctor or nurse is not listening to what you were saying when you go to a doctor's office or other health care provider? (40192394) | Never | Never (40192465) |
| | | | Rarely or Sometimes | Rarely (40192481), Sometimes (40192429) |
| | | | Always or Most of the Time | Always (40192515), Most of the Time (40192382) |
| | | | Missing | Skip (903096), No Response |
| Language Other Than English Spoken at Home | Social Factors of Health | Do you speak a language other than English at home? (40192526) | Yes | Yes (40192448) |
| | | | No | No (40192523) |
| | | | Missing | Skip(903096), Prefer Not to Answer (903079), No response |
| General Quality of Life | Overall Health | In general, would you say your quality of life is: (1585717) | Excellent | Excellent (1585718) |
| | | | Very Good | Very Good (1585719) |
| | | | Good or Fair | Good (1585720), Fair (1585721) |
| | | | Poor | Poor (1585722) |
| | | | Missing | Skip (903096), No Response |
| General Mental Health | Overall Health | In general, how would you rate your mental health, including your mood and your ability to think? (1585729) | Excellent | Excellent (903626), Excllent (1585730) |
| | | | Very Good | Very Good (1585731) |
| | | | Good or Fair | Good (1585732), Fair (1585733) |
| | | | Poor | Poor (1585734) |
| | | | Missing | Skip (903096), No response |
| General Physical Health | Overall Health | | Excellent | Excellent (1585724) |

| | | | | |
|---|---|---|---|---|
| | | In general, how would you rate your physical health? (1585723) | Very Good | Very Good (1585725) |
| | | | Good or Fair | Good (1585726), Fair (1585727) |
| | | | Poor | Poor (1585728) |
| | | | Missing | Skip (903096), No Response |
| General Health | Overall Health | In general, would you say your health is: (1585711) | Excellent | Excellent (1585712) |
| | | | Very Good | Very Good (1585713) |
| | | | Good or Fair | Good (1585714), Fair (1585715) |
| | | | Poor | Poor (1585716) |
| | | | Missing | Skip (903096), No Response |
| Cigarettes | Lifestyle | Q1: Have you smoked at least 100 cigarettes in your entire life? (There are 20 cigarettes in a pack.)? (1585857) Q2: Do you now smoke cigarettes every day, some days, or not at all? (1585860) | Never | Q1: No (1585859) |
| | | | Former | Q1: Yes (1585858) Q2: Not At All (1585863) |
| | | | Current | Q1: Yes (1585858) Q2: Every Day (1585861), Some Days (1585862) |
| | | | Missing | Otherwise |
| Vaping | Lifestyle | Q1: Have you ever used an electronic nicotine product, even one or two times? (Electronic nicotine products include e- cigarettes, vape pens, hookah pens, personal vaporizers and mods, e-cigars, e-pipes, and e-hookahs.)- (1586166) Q2: Do you now use electronic nicotine products ⋯ (1586169) | Never | Q1: No (1586168) |
| | | | Former | Q1: Yes (1586167) Q2: Not At All (1586172) |
| | | | Current | Q1: Yes (1586167) Q2: Every Day (1586170), Some Days (1586171) |
| | | | Missing | Otherwise |
| Alcohol | Lifestyle | Q1: In your entire life, have you had at least 1 drink of any kind of alcohol, not counting small tastes or | Never | Q1: No (1586200) |
| | | | Former | Q1: Yes (1586199) Q2: Never (1586202) |
| | | | Current | Q1: Yes (1586199) |

| | | | |
|---|---|---|---|
| | sips? (By a "drink," we mean a can or bottle of beer, a glass of wine or a wine cooler, a shot of liquor, or a mixed drink with liquor in it.) (1586198)<br>Q2: How often did you have a drink containing alcohol in the past year? (1586201) | | Q2: Monthly or Less (1586203), 2 to 4 Per Month (1586204), 2 to 3 Per Week (1586205), 4 or More Per Week (1586206) |
| | | Missing | Otherwise |
